# ANALYSIS OF QUALITY OF LIFE IN PATIENTS WITH CHRONIC KIDNEY DISEASE ON HEMODIALYSIS IN THE SOUTH OF SANTA CATARINA

**DOI:** 10.64898/2026.11.17.26368318

**Authors:** Vittório Augusto Cimolim, Pedro Lucas Zanette Selinger, Sérgio Emerson Sasso

## Abstract

**Introduction:** Chronic Kidney Disease (CKD) is a clinical condition influenced by several socioeconomic, demographic and anthropometric factors, especially if patients are on hemodialysis (HD).

**Objective:** Analyze the QoL in patients with chronic kidney disease on hemodialysis.

**Methodology:** The study was developed with 52 patients with chronic kidney disease undergoing hemodialysis at the SC LTDA Nephrology Clinic, at the Hospital Regional of Araranguá, Santa Catarina. The patients’ quality of life was quantified in domains obtained through the KDQOL-SF™ 1.3 questionnaire, which analyzes several aspects, including: impacts of the disease on physical and mental health, impact of the disease on daily activities and satisfaction with treatment. Another questionnaire, authored by the researchers, containing socioeconomic, demographic and anthropometric data was also applied. Finally, the results of the two questionnaires were related to see which factors influence QoL and how.

## INTRODUCTION

CKD is a pathological condition characterized by irreversibility, gradual progression, and high prevalence, affecting approximately 10–13% of the global population.^1^ It represents a growing public health concern, with both its prevalence and incidence having nearly doubled over the past three decades.^2^

CKD is a clinical syndrome resulting from a significant alteration in kidney structure or function.^1^ The disease is defined by the presence, for at least three months, of either a glomerular filtration rate (GFR) <60 mL/min/1.73 m^2^ or a GFR ≥60 mL/min/1.73 m^2^ accompanied by at least one marker of kidney damage, including an albumin excretion rate ≥30 mg/24 h, previous kidney transplantation, imaging abnormalities of the kidneys, histological abnormalities, or electrolyte disturbances secondary to tubular dysfunction.^3^

Diabetes mellitus and hypertension are among the leading causes of CKD and, together with sociodemographic factors such as low educational attainment, are associated with an increased risk of developing the disease.□ Furthermore, CKD is associated with an increased risk of cardiovascular disease, as well as greater disease severity and higher mortality rates.□In the early stages of CKD, patients are generally asymptomatic. However, as the disease progresses to kidney failure, several nonspecific symptoms may develop, including fatigue, loss of appetite, confusion, insomnia, and pruritus, in addition to complications such as anemia and mineral and bone disorders. □ □ □

CKD is typically an insidious disease that may remain clinically silent for many years before progressing to kidney failure, requiring kidney replacement therapy through dialysis or kidney transplantation. □ □ □In kidney failure, hemodialysis (HD) is one of the treatment modalities that replaces kidney function by removing metabolic waste products and excess body water while maintaining acid–base and electrolyte balance. Consequently, HD is the most commonly used therapy for kidney failure refractory to conventional medical management and is also indicated in selected cases of acute intoxication and as a bridge to kidney transplantation. □

Quality of life (QoL) is a multidimensional concept that is difficult to define, encompassing different levels of assessment, ranging from social and community well-being to the evaluation of specific circumstances affecting individuals or groups.^1^□Health-related QoL in patients with CKD is substantially impaired regardless of disease stage, as several factors commonly present in this population negatively influence this outcome, including depression, anxiety, physical inactivity, and lack of social support.^11^ In this context, CKD creates a setting that is highly conducive to deterioration in QoL, since, in addition to the fear and anxiety associated with the diagnosis itself, disease-related symptoms and medication-related adverse effects further compromise patients’ well-being and limit their ability to perform daily activities. □

Given the high prevalence, increasing incidence, and progressive nature of CKD, many patients are likely to progress to kidney failure and require HD. Therefore, identifying the factors that influence QoL among patients undergoing HD and understanding how these factors affect their well-being are of considerable clinical importance, as they may contribute to a better understanding of disease progression and outcomes. Such knowledge may also support the development of more appropriate management strategies for this patient population. Accordingly, the aim of the present study was to evaluate QoL in patients with CKD undergoing HD.

## MATERIALS AND METHODS

### Ethical considerations

The study was approved by the Research Ethics Committee (REC) of the University of Southern Santa Catarina (UNESC) under approval number 6,582,011. All participants were enrolled only after providing written informed consent.

### Study design

Analytical cross-sectional study with a quantitative approach based on primary data collection.

### Study population

Patients with dialysis-dependent CKD receiving treatment at Clínica de Nefrologia SC LTDA, Regional Hospital of Araranguá, Santa Catarina, Brazil, between March and July 2024.

### Inclusion criteria

Adults (≥18 years of age) who provided written informed consent.

### Exclusion criteria

Individuals with evident cognitive impairment.

### Sample

The final sample consisted of 52 participants after the exclusion of three patients according to the predefined eligibility criteria.

### Data Collection

Patient assessment was conducted through face-to-face interviews using a validated printed questionnaire administered by the researchers, with participants providing oral responses. The questionnaire used was the KDQOL-SF™ version 1.3.^12^ This instrument was specifically developed for patients undergoing dialysis and consists of 24 questions with their respective subscales, assessing multiple aspects of QoL and generating a final numerical score. Initially, the questionnaire generates a raw numerical value for each item. These raw scores are then transformed to a standardized scale ranging from 0 to 100, representing the percentage of the maximum attainable score. Higher scores indicate better QoL. Subsequently, the transformed scores of related items are grouped into domains according to the questionnaire manual. Finally, the mean of the transformed scores within each domain is calculated to obtain the final score for that domain.

In addition to the information obtained through the KDQOL-SF™, a second questionnaire developed by the investigators was administered. This questionnaire collected the following information: sex, age (completed years), skin color, time since CKD diagnosis (months), duration of dialysis treatment (months), educational level, presence of other chronic diseases (if present, which conditions), monthly income (number of minimum wages), weight (kg), height (cm), body mass index (BMI), calculated as weight divided by height squared (kg/m^2^), smoking status (for current or former smokers, smoking exposure was calculated in pack-years using the formula: packs smoked per day multiplied by the number of years of smoking), travel time to the dialysis center, mode of transportation used to reach the dialysis center, and physical activity practice (for physically active individuals, weekly duration of physical activity was recorded).

The questionnaire generates a total of 19 domains. The domains selected for the present study were symptoms, general health, emotional well-being, effects of kidney disease, pain, sleep, quality of social interaction, physical functioning, and energy.

The entire scoring process was based on the *KDQOL-SF™ Version 1*.*3 User’s Manual*. https://www.rand.org/content/dam/rand/pubs/papers/2006/P7994.pdf.

### Statistical Analysis

The collected data were analyzed using IBM Statistical Package for the Social Sciences (SPSS), version 23.0. Quantitative variables were expressed as median (minimum and maximum values) and as mean ± standard deviation (SD). Qualitative variables were expressed as frequencies and percentages.

Statistical analyses were performed using a significance level of α = 0.05, corresponding to a 95% confidence level. Data normality was assessed using the Shapiro–Wilk test. The homogeneity of variances for quantitative variables across categories of qualitative variables was evaluated using Levene’s test.

Comparisons of the mean values of quantitative variables between categories of dichotomous qualitative variables were performed using the independent-samples Student’s *t*-test when data followed a normal distribution, and the Mann–Whitney *U* test when the assumption of normality was not met.

Comparisons of the mean values of quantitative variables among categories of polytomous qualitative variables (when a normal distribution was observed) were performed using one-way analysis of variance (ANOVA), followed by Tukey’s post hoc test when statistically significant differences were identified.

## RESULTS

Tables 1 and 2 present the socioeconomic, demographic, and anthropometric characteristics of the 52 patients undergoing HD at the Clínica de Nefrologia LTDA of the Regional Hospital of Araranguá, Santa Catarina, between March and July 2024.

**Table 1.** Socioeconomic, demographic, and anthropometric characteristics of patients undergoing hemodialysis at a medium-complexity hospital in Southern Santa Catarina, Brazil, between March and July 2024.

|  | Mean ± SD, Median (Min. – Max.), n (%)<br>n = 52 |
|---|---|
| Age (Years) | 56,40 ± 15,51 |
| Sex |  |
| Male | 37 (71,2) |
| Female | 15 (28,8) |
| Educational level |  |
| Illiterate | 2 (3,8) |
| Incomplete elementary education | 26 (50,0) |
| Complete elementary education | 1 (1,9) |
| Incomplete high school | 3 (5,8) |
| Complete high school | 14 (26,9) |
| Incomplete Higher education | 2 (3,8) |
| Complete Higher education | 4 (7,7) |
| Race/Ethnicity |  |
| White | 38 (73,1) |
| Mixed race | 8 (15,4) |
| Black | 6 (11,5) |
| Monthly income |  |
| <1 minimum wage | 6 (11,5) |
| 1-2 minimum wages | 32 (61,5) |
| 2-3 minimum wages | 7 (13,5) |
| 3-4 minimum wages | 5 (9,6) |
| >4 minimum wages | 2 (3,8) |
| Duration of CKD diagnosis (months) | 46,5 (3,0 – 348,0) |
| Duration of hemodialysis (months) | 24,0 (0,25 – 180,0) |
| Presence of comorbidities |  |
| Yes | 44 (84,6) |
| DM | 18 (34,6) |
| Hypertension | 34 (65,4) |
| Hypothyroidism | 3 (5,8) |
| Dyslipidemia | 4 (7,7) |
| Psychiatric disorders | 6 (11,5) |
| Other | 9 (17,3) |
| No | 8 (15,4) |

SD: Standard deviation. CKD: Chronic kidney disease. DM: Diabetes Mellitus. Min: minimum. Max: maximum
Source: study data, 2024.

**Table 2.**
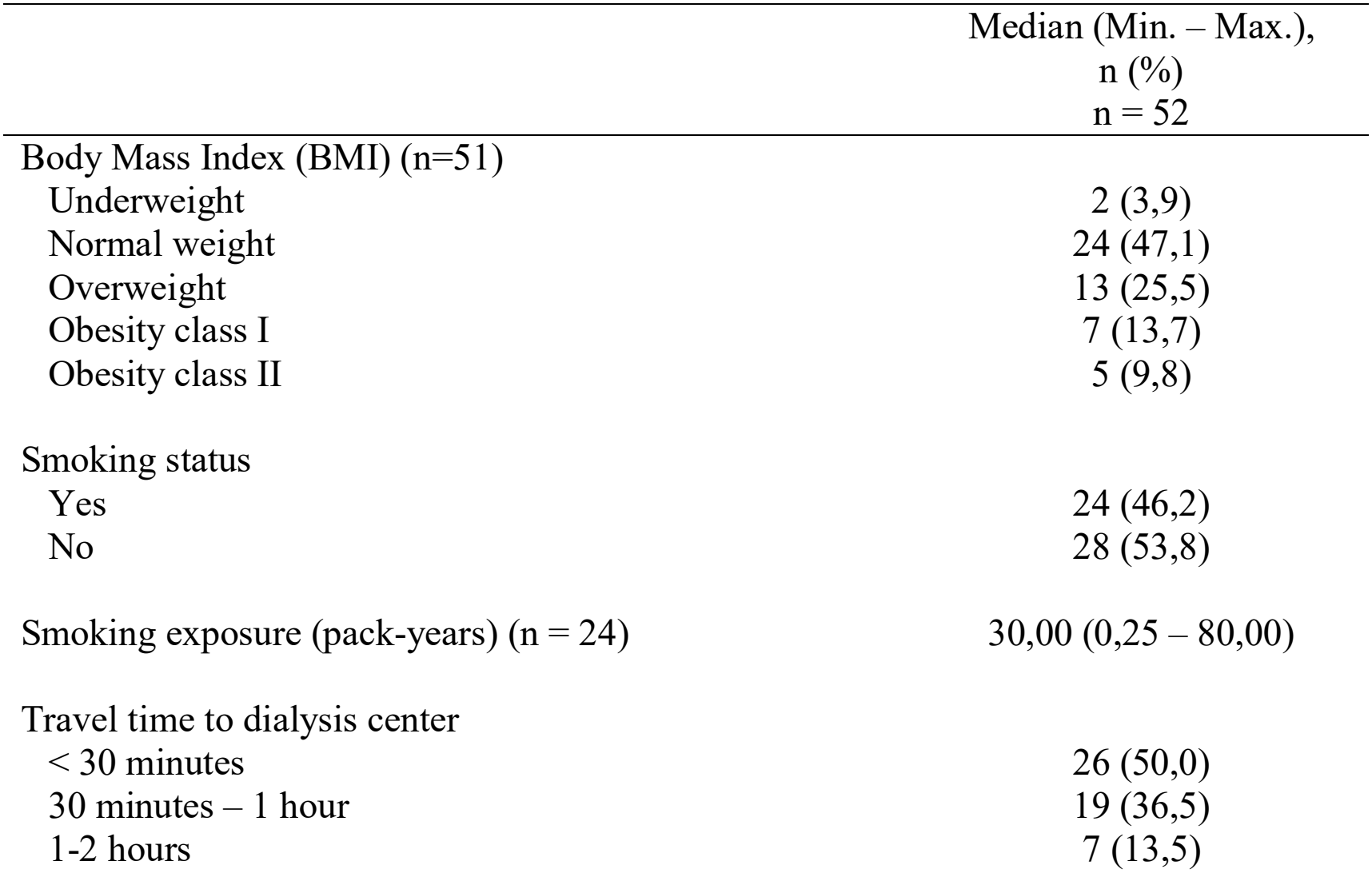
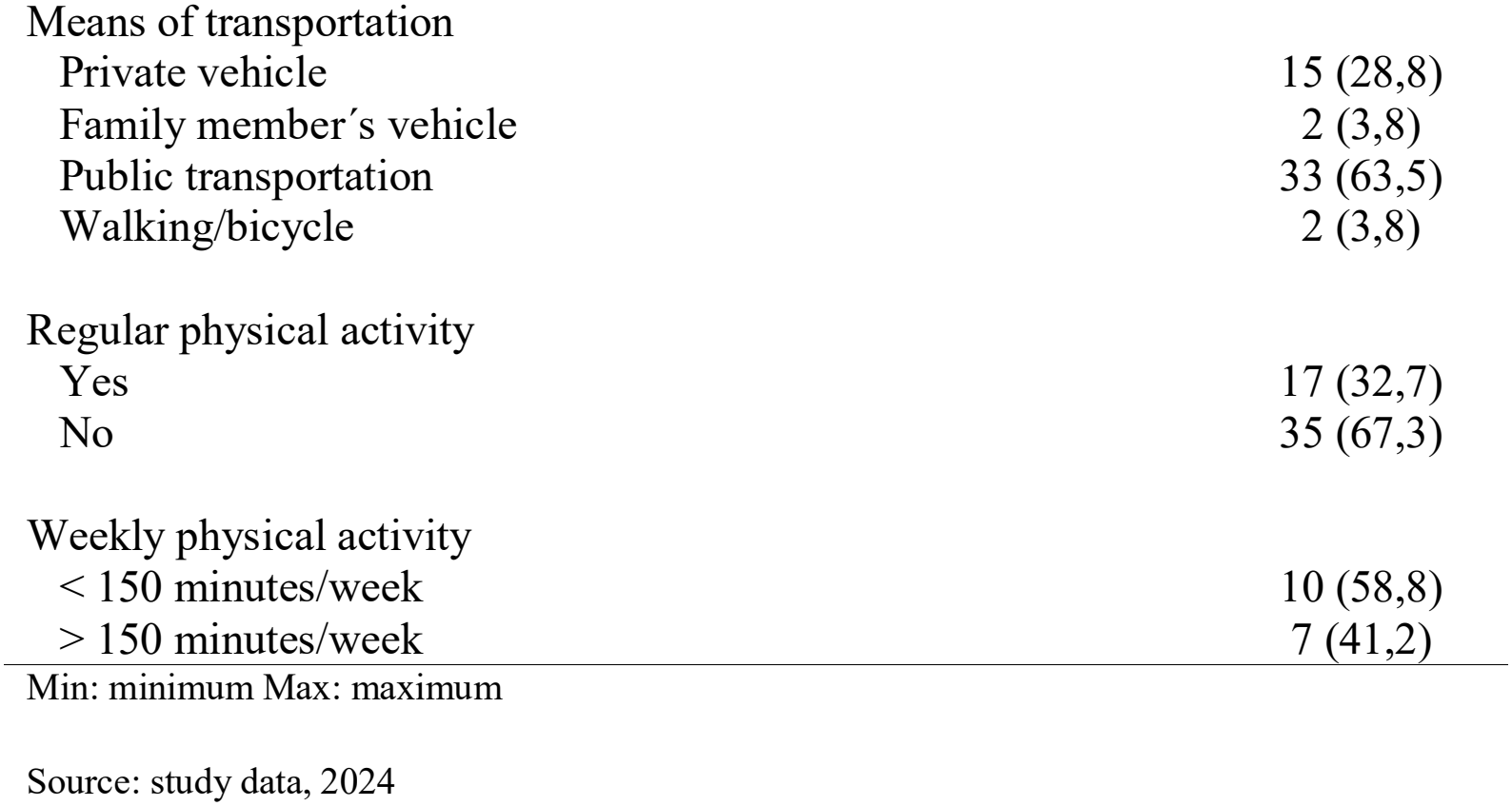
Additional socioeconomic, demographic, and anthropometric characteristics of patients undergoing maintenance hemodialysis at a medium-complexity hospital in Southern Santa Catarina, Brazil, between March and July 2024.

Table 1 shows that the mean age of the patients included in the study was 56.4 years, with 71.2% being male. Regarding educational level, 50.0% had not completed elementary school, while 26.9% had completed high school. Concerning skin color, most patients (73.1%) were White. With respect to monthly income, 61.5% earned between one and two minimum wages, whereas 13.5% earned between two and three minimum wages. The mean time since CKD diagnosis was 46.5 months, ranging from 3 to 348 months. The mean duration of dialysis treatment was 24 months, with a range of 0.25 to 180 months. Among the study participants, 84.6% had chronic diseases concomitant with CKD, of whom 65.4% had systemic arterial hypertension (SAH) and 34.6% had diabetes mellitus (DM).

According to BMI classification (Table 2), 47.1% of the patients had normal weight, 25.5% were overweight, and 13.7% had class I obesity. Regarding smoking status, 46.2% of the patients were current or former smokers, with a mean smoking exposure of 30 pack-years. Regarding travel time to the dialysis center, 50.0% of the patients required less than 30 minutes to reach the facility, whereas 36.5% required between 30 minutes and one hour. Concerning transportation, most patients (63.5%) traveled to the dialysis center using public transportation. Among the interviewed individuals, 32.7% reported engaging in physical activity, and of these, 58.8% performed less than 150 minutes of physical activity per week.

Table 3 presents all QoL domains based on the results of the KDQOL-SF™ 1.3 questionnaire. Among these, only the symptom, general health, emotional well-being, effects of kidney disease, pain, sleep, quality of social interaction, physical functioning, and energy domains were included in the present study. Among these domains, quality of social interaction showed the highest mean score (75.26; SD: 23.44), whereas general health had the lowest mean score (45.67; SD: 23.45).

**Table 3.**
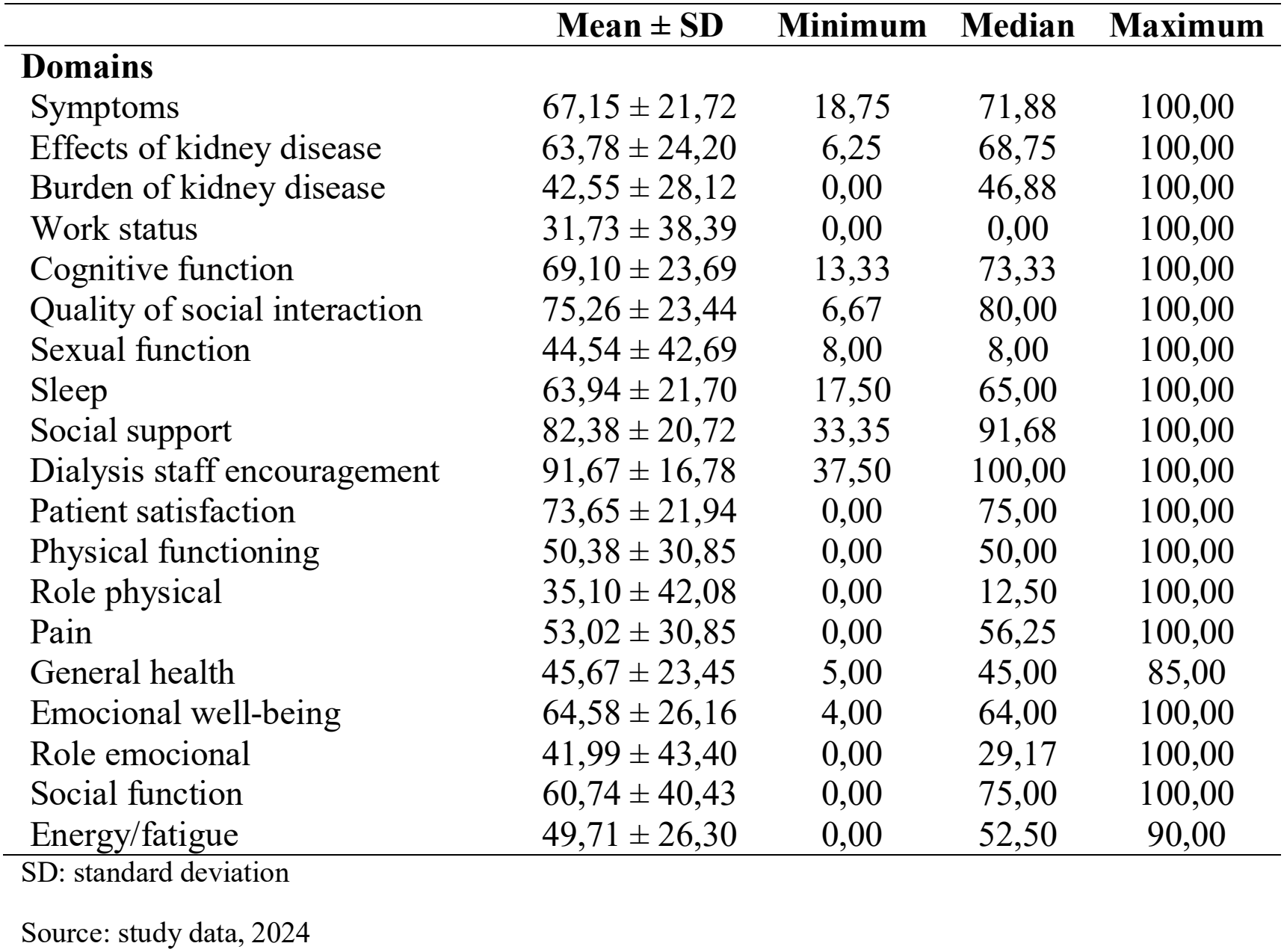
KDQOL-SF™ version 1.3 quality-of-life domain scores among patients undergoing maintenance hemodialysis at a medium-complexity hospital in Southern Santa Catarina, Brazil, between March and July 2024.

Tables 4 and 5 present the statistically significant associations between the dependent and independent variables. In Table 4, the mean symptom domain score was lower among patients with DM than among those without this comorbidity (p = 0.004). Patients with a monthly income below one minimum wage had lower emotional well-being scores than those with higher monthly incomes (p = 0.002). Patients with psychiatric disorders had lower mean scores in the emotional well-being domain than those without psychiatric disorders (p = 0.028). Female patients had lower mean scores in the effects of kidney disease domain compared with male patients (p = 0.045). In the pain domain, women also had lower mean scores than men (p = 0.003). Likewise, the mean sleep domain score was lower among women than among men (p = 0.049).

**Table 4.** Associations between clinical and epidemiological characteristics and KDQOL-SF™ version 1.3 quality-of-life domains among patients undergoing maintenance hemodialysis at a medium-complexity hospital in Southern Santa Catarina, Brazil, between March and July 2024.

|  | **Domain** | Value – p |
|---|---|---|
|  | **Symptoms** |  |
| Diabetes Mellitus |  |  |
| Yes | 55,44 ± 20,87 | 0,004<sup>†</sup> |
| No | 73,34 ± 19,76 |  |
|  | **Emocional well-being** |  |
| Monthly income |  |  |
| <1 minimum wage | 34,00 ± 21,73<sup>a</sup> | 0,002<sup>‡</sup> |
| 1-2 minimum wages | 65,25 ± 23,47<sup>b</sup> |  |
| 2-3 minimum wages | 76,57 ± 25,13<sup>b</sup> |  |
| 3-4 minimum wages | 86,80 ± 14,94<sup>b</sup> |  |
| >4 minimum wages* | 48,00 ± 33,94 |  |
| Psychiatric disorder |  |  |
| Yes | 43,33 ± 26,10 | 0,028<sup>†</sup> |
| No | 67,35 ± 25,14 |  |
|  | **Effects of kidney disease** |  |
| Sex |  |  |
| Feminine | 53,34 ± 23,70 | 0,045<sup>¥</sup> |
| Masculine | 68,14 ± 23,36 |  |
|  | **Pain** |  |
| Sex |  |  |
| Feminine | 33,50 ± 29,14 | 0,003<sup>¥</sup> |
| Masculine | 60,93 ± 28,20 |  |
|  | **Sleep** |  |
| Sex |  |  |
| Feminine | 54,67 ± 23,41 | 0,049<sup>¥</sup> |
| Masculine | 67,70 ± 20,09 |  |

<sup>†</sup> value obtained by Mann-Whitney U test;
<sup>‡</sup> value obtained by One-way ANOVA;
<sup>¥</sup> value obtained by Independent-samples Student's t-test;
<sup>a,b</sup> Different superscript letters indicate statistically significant differences according to Tukey's post hoc test (p ≤ 0.05).
\*Values were not included in the comparison because the sample size was insufficient for hypothesis testing.
Source: study data, 2024

**Table 5.**
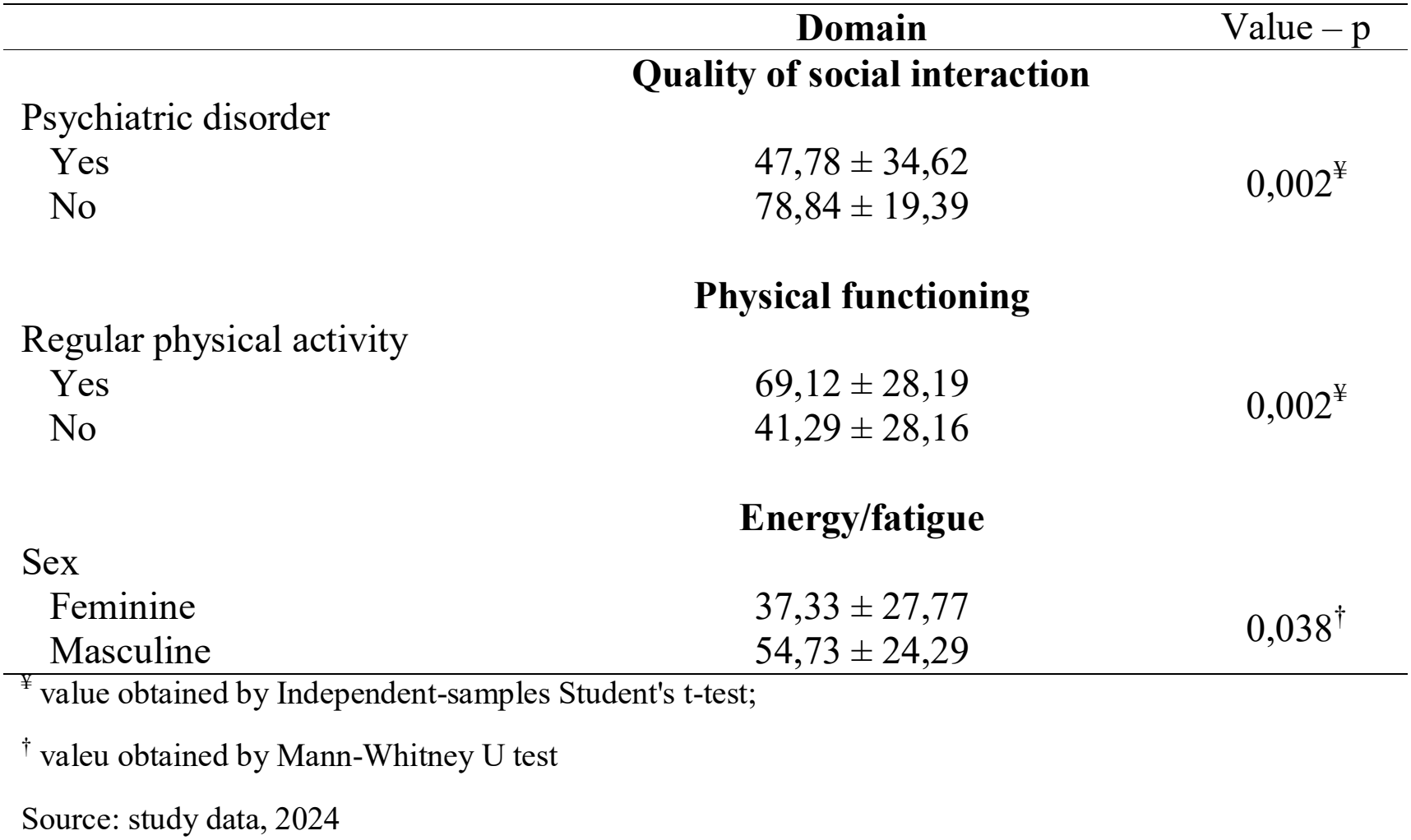
Associations between clinical and epidemiological characteristics and KDQOL-SF™ version 1.3 quality-of-life domains among patients undergoing maintenance hemodialysis at a medium-complexity hospital in Southern Santa Catarina, Brazil, between March and July 2024.

|  | **Domain** | Value – p |
|---|---|---|
|  | **Quality of social interaction** |  |
| Psychiatric disorder |  |  |
| Yes | 47,78 ± 34,62 | 0,002<sup>¥</sup> |
| No | 78,84 ± 19,39 |  |
|  | **Physical functioning** |  |
| Regular physical activity |  |  |
| Yes | 69,12 ± 28,19 | 0,002<sup>¥</sup> |
| No | 41,29 ± 28,16 |  |
|  | **Energy/fatigue** |  |
| Sex |  |  |
| Feminine | 37,33 ± 27,77 | 0,038<sup>†</sup> |
| Masculine | 54,73 ± 24,29 |  |

<sup>¥</sup> value obtained by Independent-samples Student's t-test;
<sup>†</sup> valeu obtained by Mann-Whitney U test
Source: study data, 2024

As shown in Table 5, patients with psychiatric disorders had lower scores in the quality of social interaction domain than those without psychiatric disorders (p = 0.002). Individuals who did not engage in physical activity had lower physical functioning scores than those who were physically active (p = 0.002). In the energy domain, women had lower mean scores than men (p = 0.038).

The general health domain was not significantly associated with any of the independent variables analyzed.

## DISCUSSION

As CKD is a disease with high prevalence and morbidity, when analyzing the QoL of dialysis patients, it is noteworthy that several factors influence it.^1^ □ ^11^

In the present study, DM was found to contribute to a greater symptom burden among patients undergoing HD, as evidenced by lower scores in the symptom domain. DM is one of the leading causes of CKD and is closely associated with the development of several complications, particularly in individuals with long-standing disease.

Symptoms such as foot ulcers, vision loss, and sexual dysfunction are highly prevalent among these patients.^13^ □ □ Furthermore, DM itself has been reported to negatively affect QoL, especially in the physical and social functioning domains.^1^ □ Therefore, it is plausible that both diseases reinforce one another, increasing the level of symptoms reported by patients and compromising their QoL.

Moreover, in this study, emotional well-being was found to be influenced by monthly income, as patients earning less than one minimum wage scored lower in this domain than those with higher monthly incomes. Consistent with this finding, the literature reports that monthly income and well-being are closely related and directly proportional, with one increasing as the other increases.^1^ □ □ ^1^ □ In this context, one possible explanation for this issue is that lower purchasing power may be associated with reduced access to healthcare services, such as medical consultations, medications, and adjunctive therapies, which are considered essential for maintaining well-being in individuals with chronic diseases.^1^ □

Furthermore, this study demonstrated that individuals with psychiatric disorders presented lower scores in the emotional well-being and quality of social interaction domains. Mental health is known to be a major contributor and an essential component of well-being, and when it is impaired, well-being is inevitably affected as well.^1^ □ This issue is of exceptional importance, given that psychiatric disorders are highly prevalent and constitute the leading cause of global burden on healthcare services.^1^ □ In addition, CKD itself has been associated with the development of cognitive decline and psychiatric disorders, creating a vicious and harmful cycle that reduces patients’ well-being and, consequently, their QoL.^2^ □ □ ^21^

In the present study, both the effects of kidney disease and patient-reported pain were more frequently reported by women than by men. The effects of CKD may be described as the impact of the disease on daily life, including dietary restrictions, work and travel limitations, stress, and sexual life.^12^ In this regard, studies have shown that women tend to report more pain and other complaints than men in response to the same stimulus or insult, which may explain their lower scores.^22^ Additionally, there is a cultural aspect related to the male role, whereby many men perceive masculinity as synonymous with strength and resilience in the face of adversity. Consequently, many do not report their pain and symptoms even when they are present.^23^

In this study, women reported poorer sleep quality compared with men. First, there is evidence that sleep disorders in general are more common among individuals with CKD than in the general population, given the symptoms and challenges imposed by the disease.^2^ □ Moreover, healthy women, particularly younger women, have been shown to have a 28% higher risk of insomnia than men, with this risk increasing further among those in the perimenopausal period.^2^ □ □ ^2^ □ The reason for poorer sleep among women of reproductive age remains unclear. Some theories suggest that female hormones (estrogen and progesterone) contribute to sleep disturbances; however, this finding remains controversial and requires further investigation.^2^ □ Among perimenopausal women, sleep deterioration is generally attributed to menopausal symptoms such as hot flashes and mood changes.^2^ □ □ ^2^ □

In the current study, lack of physical activity negatively affected the physical functioning domain. It is well established that physical activity improves physical capacity, enabling individuals to perform more demanding activities with fewer difficulties, which explains the reduced limitations in daily life among those who engage in exercise.^3^ □ Furthermore, physical exercise has been shown to be closely associated with physical functioning, as studies have demonstrated that, particularly among women, those who engage in greater levels of physical activity exhibit better physical functioning.^31^ Finally, physical activity not only improves specific domains but has also been associated with an overall improvement in QoL.^32^

This study also found that women reported lower energy levels than men. In this regard, even among healthy individuals, the incidence of fatigue tends to be higher in women than in men, with childbirth and socially imposed roles being considered possible factors contributing to the vulnerability of this group.^33^ Furthermore, potential biomarkers associated with fatigue have been identified and are particularly common among working women, suggesting the existence of a biological component underlying this issue.^3^ □ However, it is important to note that this factor has not been associated with a worse prognosis among women with CKD, as men are the group that experiences a more rapid decline in GFR and a higher rate of pre-dialysis mortality.^3^ □

## CONCLUSION

This study aimed to evaluate the factors associated with QoL in patients with CKD undergoing HD. We concluded that, among the QoL domains, quality of social interaction showed the highest scores, whereas general health was the most negatively affected domain. We also found that individuals with DM experienced a greater symptom burden compared with other patients. Furthermore, emotional well-being was lower among patients with a monthly income below one minimum wage, and individuals with psychiatric disorders exhibited greater impairment in both emotional well-being and quality of social interaction. It was also concluded that the lack of physical activity negatively affects the QoL of these patients. Additionally, women undergoing dialysis for kidney disease experience a greater impact of CKD across multiple aspects of QoL.

As improving QoL is the primary goal of all medical interventions, understanding the factors that influence it is essential, including among patients with dialysis-dependent CKD, given that this condition is, in most cases, incurable.

Therefore, routine assessment of QoL in patients undergoing dialysis is recommended.

## Data Availability

All data produced in the present work are contained in the manuscript

https://www.rand.org/content/dam/rand/pubs/papers/2006/P7994.pdf.

## ACKNOWLEDGMENTS

The authors sincerely thank all patients who agreed to participate in this study. We are also grateful to the hemodialysis staff of the SC LTDA Nephrology Clinic for their cooperation, hospitality, and support throughout the data collection process.

Finally, we express our special appreciation to Érica Sipriano for her valuable assistance with the statistical analyses performed in this study.

